# Quality of life in head and neck cancer survivors: the Big Data for Quality of Life study

**DOI:** 10.1101/2024.02.02.24302118

**Authors:** Mauricio Moreira-Soares, Erlend I. F. Fossen, Katherine J. Taylor, Susanne Singer, Katrina Hurley, Steve Thomas, Miranda Pring, Andrew Ness, Stefano Cavalieri, Claudia Vener, Laura Lopez-Perez, Maria Fernanda Cabrera-Umpierrez, Giuseppe Fico, Arnoldo Frigessi, Lisa Licitra, Marissa LeBlanc, BD4QoL consortium

## Abstract

**Background:** The Big Data for Quality of Life (BD4QoL) study investigates quality of life (QoL) in head and neck cancer (HNC) survivors, focusing on survivorship and characterizing survivor demographics.

**Methods:** We screened data from 5 studies across Europe (N=7276) and included patients with a diagnosis of squamous cell carcinoma (oral cavity, hypopharynx, larynx, oropharynx, nasal cavity and paranasal sinuses), treated with curative intent, alive after treatment, TNM 7^th^ ed. stages I, II, III, IVa and IVb, with availability of QoL questionnaires.

**Results:** The cohort of 4448 HNC survivors primarily includes men (78%) with median age 61 years. Most received radiotherapy (75%) and had a history of smoking (78%). Survivors’ scores on EORTC QLQ-C30 functioning scales indicated high functioning, with prevalent symptoms of fatigue, pain, and insomnia. Lower rates of missing data were observed in older patients, those with higher education and income levels, nonsmokers, married individuals, and patients not treated with radiotherapy. The odds ratios ranged from 0.47 to 0.99, indicating these factors may predict more consistent QoL data reporting in HNC survivors.

**Conclusions:** These data support the development and validation of clinical prediction models for QoL in HNC survivors in a multicentre randomized controlled trial.

## 1 Introduction

The incidence of head and neck cancer (HNC) has been increasing worldwide in the last years and reached 1.1 million new diagnoses in 2016^1,2^. Globally, it is the seventh most common type of cancer^3,4^. At the same time, the overall 5-year survival has improved considerably in the last decades, changing from 55% in 1992-1996 to 66% in 2002-2006^5^, which makes long-term quality of life (QoL) a key concern for patients. These changes in survival can be partially explained by better treatments and a deeper understanding of the disease mechanisms, but also due to an increasing proportion of human papilloma virus (HPV)-induced oropharyngeal tumours which mainly affect a younger population with fewer comorbidities and better prognosis^6–10^. According to the stage of the disease and patient status (i.e., performance status, comorbidities), HNC can be treated with either single modality or multimodal approaches, including combinations of surgery, radiotherapy, and systemic therapy^11^. Despite the intention to cure, recurrences in the local and regional areas, as well as distant relapses, are common^12,13^. Additionally, these treatment methods often lead to significant toxicities and long-term complications^14–16^. Consequently, the QoL of HNC survivors is frequently compromised. Health related QoL has been found to be associated with clinical endpoints in oncology patients^17^, particularly studies have shown strong evidence of association between physical functioning and global QoL change with overall survival in individuals with HNC^18^. However, there is limited research on the long-term changes in QoL and the factors that influence these changes. Existing evidence suggests that global QoL tends to recover within 12 months after HNC treatment, but late complications persist, including declines in physical functioning, fatigue, xerostomia (dry mouth), and sticky saliva^19^, affecting overall QoL. Furthermore, the available literature on QoL in HNC survivors is relatively limited, particularly concerning long-term changes and determinants of QoL over time^20^. Most studies have focused on short-term recovery, but there is a lack of information regarding the sustained effects and late sequelae experienced by HNC survivors^21,22^. Investigating these factors is essential for optimizing patient care, identifying potential interventions to alleviate specific needs, and improving survivorship outcomes.

This study describes the creation of the multi-national Big Data for Quality of Life (BD4QoL) historical cohort, which was established to investigate QoL in HNC survivors. This cohort will be used for research to better understand the QoL trajectory in HNC survivors.

The aim of the study is to define and describe clinical, demographic, quality of life and behavioural characteristics of the patients in the BD4QoL historical cohort.

## 2 Methods

### 2.1 Quality of life questionnaires

The EORTC QLQ-C30 is a questionnaire developed by the European Organization for Research and Treatment of Cancer, for assessing the quality of life of cancer patients ^23,24^. This questionnaire is a Patient Reported Outcome (PRO) instrument which contains 30 questions that compose 10 sub-scales divided in three groups: functional sub-scales (physical function, role function, cognitive function, emotional function and social function), symptom sub-scales (pain, fatigue, nausea/vomiting) and global health status (GHS)/quality of life. In addition, it contains 6 individual items to assess: dyspnoea, insomnia, appetite loss, constipation, diarrhoea and financial difficulties. All scales and single-item measures range in score from 0 to 100. A high scale score represents a higher response level. Thus, a high score for a functional scale represents a high/healthy level of functioning, a high score for GHS/QoL represents a good overall QoL, but a high score for a symptom scale/item represents a high level of symptoms/problems.

The head and neck cancer module (EORTC QLQ-H&N35) incorporates seven multi-item scales that assess pain, swallowing, senses (taste and smell), speech, social eating, social contact and sexuality. There are also eleven single items. For all items and scales, high scores indicate a higher degree of problems, i.e., there are no functioning scales.

The EORTC QLQ-HN43 module is a revised and updated version of the head and neck cancer module EORTC QLQ-HN35. The 43 items can be combined into the following scales: fear of progression, body image, dry mouth and sticky saliva, pain in the mouth, sexuality, problems with senses, problems with shoulder, skin problems, social eating, speech, swallowing, and problems with teeth. Single item scales are coughing, swelling in the neck, neurological problems, trismus, social contact, weight loss, and problems with wound healing.

### 2.2 Cohort participants

#### 2.2.1 Datasets

We screened data collected in prior research projects at Istituto Nazionale dei Tumori (Italy), University Hospitals Bristol and Weston NHS Foundation Trust, and University of Bristol (UK) and University Medical Centre Mainz (UMM) in Germany to construct this cohort (Table 1). Data collection was performed with the understanding and written consent of patients enrolled in the original studies.

**Table 1.** Historical datasets included in BD4QoL. Study ID is the ID used in this manuscript; study short name is the original short study name. Shown are the number of cases before eligibility filtering (N), the number of eligible cases according to the eligibility criteria in this study (Eligible). Eligible number of cases could not be assessed for BD2Decide because a selection was made prior to data transfer.

| Study ID | Study short name | References | Region | N | Eligible |
| --- | --- | --- | --- | --- | --- |
| **HN5000** | HN5000 | <sup>25,26</sup> | UK | 5404 | 3572 |
| **UMM1** | TLE | <sup>27</sup> | DE | 473 | 180 |
| **UMM2** | PLE | <sup>28</sup> | DE | 468 | 426 |
| **UMM3** | HN43 Phase IV | <sup>29</sup> | DE | 812 | 158 |
| **INT** | BD2Decide | <sup>30</sup> | IT | 119 | 112* |
| **Total** | **BD4QoL** |  | **EU** | **7276** | **4448** |

\*The data was filtered for QoL availability before data transfer (INT)

Head and Neck 5000 (HN5000) is a large UK based study of people with head and neck cancer^25,26^. The study is sponsored by University Hospitals Bristol and Weston NHS Foundation Trust (UHBW) and is run by UHBW and the University of Bristol. Briefly, 5511 people were recruited from 76 UK centres between 2011 and 2014 making it one of the largest prospective cohort studies of people with head and neck cancer in the world. The study collected more than 200 variables at several timepoints (from diagnosis to 3 years follow-up), including clinical and demographic characteristics, standard QoL questionnaires, and information about physical and mental health. The inclusion criteria were^25^: individuals over the age of 16 with a new head and neck primary cancer seen or discussed at an appropriate multidisciplinary team (MDT) meeting or clinic; People presenting with a cancer of unknown primary (CUP); those without a definitive histological diagnosis were eligible if the MDT decision was that the primary site was likely to be a HNC. The exclusion criteria included: people considered to meet the criteria for mental incapacity or vulnerability set out in the mental capacity/ vulnerable adult act, recurrent HNC, a second head and neck cancer, skin cancer, lymphoma and a histological diagnosis of Carcinoma in Situ with no clear evidence of invasion (these patients were eligible if later upstaged following MDT discussion); Patients who had already commenced their cancer treatment (with the exception of those whose treatment was also their diagnostic procedure) were also excluded.

The second data set (UMM1)^27^ was a prospective cohort study in patients before and after total laryngectomy (TLE). Further eligibility criteria were written informed consent and age of 18 years or older. The patients were interviewed in a face-to-face setting before the surgery (t1), shortly before discharge from the hospital (t2), at the end of rehabilitation (t3), one year after baseline (t4) as well as two (t5) and three years (t6) after baseline. Participants also completed self-administered questionnaires, including the EORTC QLQ-C30 and the EORTC QLQ-H&N35. A total of 389 patients were enrolled between the years 2001-2011 from 13 hospitals in Germany.

The third data set (UMM2)^28^ comes from a similar study, but this time in patients who were scheduled for partial laryngectomy (PLE). The study design and data collection were in parallel with the UMM1 study up to t4. Data collection began in 2007 and ended in 2015. A sample of 391 patients were enrolled from 16 hospitals in Germany.

The fourth data set (UMM3)^29^ comes from an international validation study for the update of the EORTC QLQ-H&N35 questionnaire, the HN43 Phase IV study. In total, 812 patients from 18 countries in Europe, the Americas and Asia were enrolled. Patients with cancer of the larynx (ICD-10 code C32), lip (C00), oral cavity (C01-06), salivary glands (C07-08), oro-hypopharynx (C09-10, C12-14), nasopharynx (C11), nasal cavity (C30), nasal sinuses (C31), sarcoma in the head and neck region (C49), and lymph node metastases from Missing primary in the head and neck area (C77, C80.0).were included. There were no restrictions regarding stage, recurrence status, or treatments planned or performed. Patients with a tumour of the eyes, orbit, thyroid, skin (even if in the head and neck area), or lymphomas in the head and neck region were excluded. Patients completed the questionnaires up to 14 days before start of treatment (t1), three months (t2), and six months thereafter (t3).

The Big Data to Decide (BD2Decide) project was a European multicentre observation study including 1537 HNC patients from Italy, Germany and the Netherlands^30^. The aim of this project was to develop a multisource database to allow for prognostic prediction modelling in loco-regionally advanced HNC patients. The database was made of two cohorts: retrospective (diagnosis 2008-2014), and prospective (diagnosis 2015-2017). Main inclusion criteria were diagnosis of head and neck squamous cell carcinoma (HNSCC); stage III and IVA/B (based on AJCC/UICC seventh edition); receiving treatments with curative intent; availability of pre-treatment tumour specimen for biological analysis; availability of pre-treatment imaging scans for radiomic analysis; for patients enrolled prospectively, PROs were collected (EORTC C30, EORTC HN35 and EQ-5D-5L). The BD4QoL study included the prospective BD2Decide patients enrolled at one of the Italian cancer centres.

These longitudinal studies enrolled patients at diagnosis and before treatment initiation.

In the BD4QoL study, survivors are defined by the following inclusion and exclusion criteria:

##### Inclusion criteria

1. Non-metastatic head and neck cancers from one of the following subsites (ICD in annex 1): oral cavity, hypopharynx, larynx, oropharynx, nasopharynx, major or minor salivary glands, nasal cavity, paranasal sinuses.
2. Having received and concluded treatments with curative intent at time of study inclusion.
3. Being alive and disease-free at last post-treatment follow-up.
4. Stage I, II, III, IVa or IVb according to TNM 7^th^ edition^31^.
5. Age ≥18 years.

##### Exclusion criteria

1. Histologies other than squamous cell carcinoma and salivary gland carcinomas (e.g., sarcoma, melanoma are excluded). Thyroid cancers, neuroendocrine tumours and non-epithelial HNC (e.g., melanoma, sarcoma, etc.) are excluded.
2. Distant metastases at the time of study entry.
3. Any previous HNC unrelated to the primary HNC for which the participant was treated; premalignant lesions (e.g., leukoplakia, erythroplakia, lichen etc.) are allowed.
4. Subjects with previous malignancies (except localized non-melanoma skin cancers, and the following in situ cancers: bladder, gastric, colon, esophageal endometrial, cervical/dysplasia, melanoma, or breast) unless a complete remission was achieved at least 5 years prior to study entry and no additional therapy is required during the study period.

#### 2.2.2 Baseline definition

HNC survivors were defined as those patients alive and disease free after the end of treatment. However, end of treatment was not recorded in HN5000 and BD2Decide studies, and the QoL measurements were not exactly aligned with end of treatment (Table 2). Therefore, we defined the BD4QoL “baseline” as the first available QoL measurement after the known or inferred end of treatment. The baseline for UMM1 and UMM2 studies is defined as 4 months after diagnosis and for UMM3 is at 3 months, when end of treatment is recorded. For BD2Decide, the baseline is defined at 6 months after diagnosis, when all patients are assumed to have finished treatment in this study. In the case of HN5000, the baseline is established at 12 months after diagnosis, when all patients have finished their curative treatment.

**Table 2.** Planned data collection per study. All studies collected data at diagnosis/before treatment (0 months), but end of treatment varies across study. UMM studies recorded end of treatment, either with start of patient rehabilitation or hospital discharge at approximately 4 months. In BD2Decide and HN5000 study all patients are assumed to have concluded treatment before 6 and 12 months respectively.

| study/time (months) | 0 | 3 | 4 | 6 | 12 | 18 | 24 | 36 |
| --- | --- | --- | --- | --- | --- | --- | --- | --- |
| BD2Decide | x |  |  | EoT |  | x | x |  |
| UMM1 | x |  | EoT | x | x |  | x | x |
| UMM2 | x |  | EoT | x | x |  |  |  |
| UMM3 | x | EoT |  | x |  |  |  |  |
| HN5000 | x |  | x |  | EoT |  |  | x |

EoT – End of treatment

### 2.3 Data harmonization

We performed data harmonization in a subset of variables to achieve compatible and comparable measurements across the different studies following the BD4QoL ontology^32^. In total, we harmonized 29 variables, consisting of 14 demographic and clinical variables and 15 QoL domains (Table 3). In addition, survival time and status were recorded in HN5000, BD2Decide and UMM1 studies, while UMM2 and UMM3 presented interval censored data.

**Table 3.** Baseline cohort characteristics for the BD4QoL historical cohort stratified by study.

| Characteristic | INT<br>N = 112<sup>1</sup> | UMM1<br>N = 180<sup>1</sup> | UMM2<br>N = 426<sup>1</sup> | UMM3<br>N = 158<sup>1</sup> | UOB<br>N = 3572<sup>1</sup> | Overall<br>N = 4448<sup>1</sup> |
| --- | --- | --- | --- | --- | --- | --- |
| Sex, n (%) |  |  |  |  |  |  |
| 1 - Male | 85 (76) | 163 (91) | 390 (92) | 118 (75) | 2,694 (75) | 3,450 (78) |
| 2 - Female | 27 (24) | 17 (9.4) | 36 (8.5) | 40 (25) | 878 (25) | 998 (22) |
| Age at diagnosis | 59 (53, 67) | 55 (50, 64) | 63 (54, 71) | 66 (57, 73) | 61 (54, 68) | 61 (54, 68) |
| Missing | 0 | 26 | 108 | 0 | 0 | 134 |
| Education level, n (%) |  |  |  |  |  |  |
| 1 - Low | 0 (NA) | 75 (52) | 147 (45) | 54 (37) | 1,197 (47) | 1,473 (47) |
| 2 - Medium | 0 (NA) | 50 (35) | 133 (41) | 25 (17) | 886 (35) | 1,094 (35) |
| 3 - High | 0 (NA) | 19 (13) | 47 (14) | 66 (46) | 450 (18) | 582 (18) |
| Missing | 112 | 36 | 99 | 13 | 1,039 | 1,299 |
| Household income (€/month), n (%) |  |  |  |  |  |  |
| 1 - Less than 500 | 0 (NA) | 19 (17) | 8 (2.9) | 0 (NA) | 367 (16) | 394 (15) |
| 2 - 500 - 1500 | 0 (NA) | 50 (43) | 97 (36) | 0 (NA) | 696 (30) | 843 (31) |
| 3 - 1501 - 2500 | 0 (NA) | 36 (31) | 113 (42) | 0 (NA) | 480 (21) | 629 (23) |
| 4 - 2501 - 3500 | 0 (NA) | 7 (6.1) | 42 (15) | 0 (NA) | 188 (8.1) | 237 (8.8) |
| 5 - more than 3500 | 0 (NA) | 3 (2.6) | 12 (4.4) | 0 (NA) | 581 (25) | 596 (22) |
| Missing | 112 | 65 | 154 | 158 | 1,260 | 1,749 |
| Body mass index | NA | NA | NA | NA | 26 (23, 29) | 26 (23, 29) |
| Missing | 112 | 180 | 426 | 158 | 1,064 | 1,940 |
| Marital status, n (%) |  |  |  |  |  |  |
| 1 - Single | 0 (NA) | 23 (16) | 32 (9.8) | 51 (32) | 331 (12) | 437 (13) |
| 2 - Married/Living with a partner | 0 (NA) | 90 (62) | 214 (65) | 107 (68) | 1,830 (69) | 2,241 (68) |
| 3 - Divorced/Separated | 0 (NA) | 23 (16) | 58 (18) | 0 (0) | 327 (12) | 408 (12) |
| 4 - Widowed | 0 (NA) | 9 (6.2) | 23 (7.0) | 0 (0) | 177 (6.6) | 209 (6.3) |
| Missing | 112 | 35 | 99 | 0 | 907 | 1,153 |
| Smoker status at diagnosis, n (%) |  |  |  |  |  |  |
| 1 - Current | 38 (34) | 34 (24) | 74 (34) | 0 (NA) | 500 (20) | 646 (21) |
| 2 - Former | 39 (35) | 100 (69) | 121 (55) | 0 (NA) | 1,487 (58) | 1,747 (57) |
| 3 - Never | 35 (31) | 10 (6.9) | 24 (11) | 0 (NA) | 577 (23) | 646 (21) |
| Missing | 0 | 36 | 207 | 158 | 1,008 | 1,409 |
| Units of alcohol per month | 193 (129, 451) | 47 (0, 159) | 53 (5, 160) | NA | 88 (36, 172) | 84 (28, 172) |
| Missing | 80 | 40 | 209 | 158 | 1,558 | 2,045 |
| Tumour location, n (%) |  |  |  |  |  |  |
| 1 - Oral cavity | 25 (22) | 0 (0) | 0 (0) | 83 (53) | 971 (27) | 1,079 (24) |
| 2 - Oropharynx | 66 (59) | 0 (0) | 2 (0.5) | 0 (0) | 1,509 (42) | 1,577 (35) |
| 3 - Nasopharynx | 0 (0) | 0 (0) | 0 (0) | 0 (0) | 69 (1.9) | 69 (1.6) |
| 4 - Hypopharynx | 6 (5.4) | 25 (14) | 16 (3.8) | 21 (13) | 135 (3.8) | 203 (4.6) |
| 5 - Larynx | 15 (13) | 140 (78) | 390 (92) | 49 (31) | 844 (24) | 1,438 (32) |
| 6 - Nasal cavity and paranasal sinuses | 0 (0) | 0 (0) | 0 (0) | 5 (3.2) | 44 (1.2) | 49 (1.1) |
| 7 - Overlapping several areas | 0 (0) | 15 (8.3) | 18 (4.2) | 0 (0) | 0 (0) | 33 (0.7) |
| UICC 7th Ed., n (%) |  |  |  |  |  |  |
| 1 - I | 0 (0) | 5 (3.0) | 211 (53) | 34 (26) | 851 (24) | 1,101 (25) |
| 2 - II | 0 (0) | 20 (12) | 97 (24) | 23 (18) | 661 (19) | 801 (18) |
| 3 - III | 21 (19) | 45 (27) | 32 (8.1) | 20 (15) | 497 (14) | 615 (14) |
| 4 - IV | 91 (81) | 97 (58) | 57 (14) | 53 (41) | 1,554 (44) | 1,852 (42) |
| Missing | 0 | 13 | 29 | 28 | 9 | 79 |
| Surgery, n (%) |  |  |  |  |  |  |
| 1 - Yes | 39 (35) | 168 (94) | 171 (40) | 82 (55) | 1,430 (40) | 1,890 (43) |
| 2 - No | 73 (65) | 10 (5.6) | 254 (60) | 66 (45) | 2,142 (60) | 2,545 (57) |
| Missing | 0 | 2 | 1 | 10 | 0 | 13 |
| Radiotherapy, n (%) |  |  |  |  |  |  |
| 1 - Yes | 112 (100) | 115 (80) | 76 (32) | 99 (63) | 2,742 (77) | 3,144 (75) |
| 2 - No | 0 (0) | 29 (20) | 163 (68) | 59 (37) | 822 (23) | 1,073 (25) |
| Missing | 0 | 36 | 187 | 0 | 8 | 231 |
| Chemotherapy, n (%) |  |  |  |  |  |  |
| 1 - Yes | 89 (79) | 15 (20) | 29 (11) | 43 (27) | 1,520 (43) | 1,696 (41) |
| 2 - No | 23 (21) | 61 (80) | 228 (89) | 114 (73) | 2,042 (57) | 2,468 (59) |
| Missing | 0 | 104 | 169 | 1 | 10 | 284 |
| Country, n (%) |  |  |  |  |  |  |
| Brazil | 0 (0) | 0 (0) | 0 (0) | 10 (6.3) | 0 (0) | 10 (0.2) |
| Egypt | 0 (0) | 0 (0) | 0 (0) | 4 (2.5) | 0 (0) | 4 (<0.1) |
| Germany | 0 (0) | 180 (100) | 426 (100) | 14 (8.9) | 0 (0) | 620 (14) |
| Israel | 0 (0) | 0 (0) | 0 (0) | 6 (3.8) | 0 (0) | 6 (0.1) |
| Italy | 112 (100) | 0 (0) | 0 (0) | 12 (7.6) | 0 (0) | 124 (2.8) |
| Japan | 0 (0) | 0 (0) | 0 (0) | 1 (0.6) | 0 (0) | 1 (<0.1) |
| Norway | 0 (0) | 0 (0) | 0 (0) | 22 (14) | 0 (0) | 22 (0.5) |
| Poland | 0 (0) | 0 (0) | 0 (0) | 10 (6.3) | 0 (0) | 10 (0.2) |
| Portugal | 0 (0) | 0 (0) | 0 (0) | 9 (5.7) | 0 (0) | 9 (0.2) |
| Spain | 0 (0) | 0 (0) | 0 (0) | 7 (4.4) | 0 (0) | 7 (0.2) |
| Sweden | 0 (0) | 0 (0) | 0 (0) | 35 (22) | 0 (0) | 35 (0.8) |
| UK | 0 (0) | 0 (0) | 0 (0) | 21 (13) | 3,572 (100) | 3,593 (81) |
| USA | 0 (0) | 0 (0) | 0 (0) | 7 (4.4) | 0 (0) | 7 (0.2) |

<sup>1</sup>n (%); Median (IQR)

Beyond the harmonised data, the studies that contributed data to this project recorded between hundreds to more than 2000 variables, which are therefore available in subsets of the BD4QoL cohort. Two versions of the head and neck module of the EORTC questionnaire were used in the studies, in addition to demographics and several other questionnaires which attempt to capture fear of recurrence, personal costs, hospital anxiety and depression, and general health.

### 2.4 Statistical analysis

Survival curves, stratified by study, were estimated using the Kaplan-Meier estimator. Survival curves were only made for the BD2Decide, HN5000 and UMM1 studies, as other studies presented interval censored data with few measurements.

To show the trajectory of overall quality of life over time, conditional on survival, we estimated the mean GHS/QoL for each time point with a QoL measurement in each study. A 95% confidence interval for the means were obtain by bootstrap with 1000 bootstrap resamples per time point.

To explore the factors associated with missing quality of life (QoL) measurements, univariate logistic regression analyses were performed. The response variable was a binary indicator for whether the GHS/QoL scale was missing or not. This scale was chosen because it summarises global QoL. The covariates considered in the respective univariate models were age, sex, tumour stage, tumour location, treatment, income, education, smoking status, and alcohol consumption.

For interpretation of the EORTC QLQ-C30 scales we used the thresholds for clinical importance (TCIs) proposed by Giesinger et al., which allow to identify patients with clinically important problems or symptoms^33^.

## 3 Results

### 3.1 Cohort characteristics

In total, 4448 HNC survivors were eligible for inclusion in this study (Figure 1). INT contributed 112 subjects with QoL measurements under BD2Decide semantics^30,34,35^. UMM contributed 883 subjects across 3 different longitudinal cohort studies (UMM1, UMM2, and UMM3), and Bristol contributed 3587 subjects using Head and Neck 5000 semantics and documentation available online (headandneck5000.org.uk). All datasets contain quality of life scores based on the EORTC QLQ-C30 and EORTC QLQ-H&N35 or EORTC QLQ-HN43 questionnaires.

**Figure 1.**
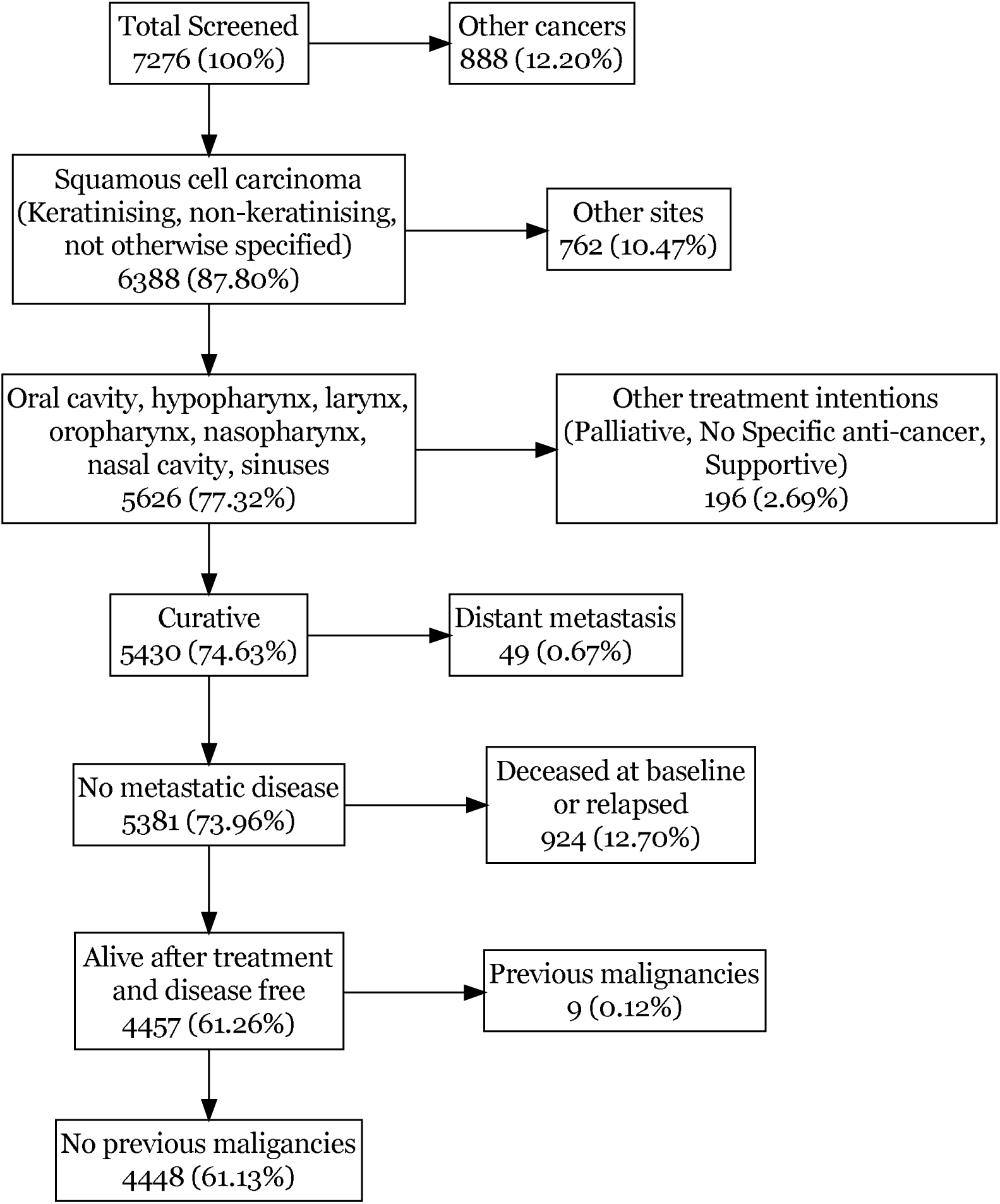
Cohort flowchart for the entire BD4QoL cohort. See Supplementary Information for flowcharts for each study.

The patients in the cohort were predominately male (78%), had a median age of 61 years, had primarily low (47%) to medium education level (35%), most were married or lived with a partner (68%), had a history of smoking (78%) and high alcohol consumption (median 84 alcohol units^1^/month) (Table 3). The education level variable presented distinct definitions across studies due to different educational systems. Thus, the education levels were mapped to years of education and converted into three categories: low (<10 years), medium (from 10 to 12 years), and high (>12 years). Overall, 75% of patients underwent radiotherapy, while 41% were treated with surgery and 38.6% received chemotherapy. Nearly half of the patients (48%) were treated with at least two therapeutic modalities, with the combination of chemotherapy and radiation being the most prevalent at 29%. The pairing of radiotherapy and surgery was the next most common, involving 10% of patients, and a combination of all three treatment approaches was used in 8% of cases. There was considerable heterogeneity between the datasets in some characteristics, particularly tumour stage and tumour region, where some studies had enrolled patients with only specific regions or tumour stages. For instance, in the BD2Decide dataset, only loco-regionally advanced stage tumours were represented (stages III and non-metastatic IV in TNM 7^th^). The UMM1 dataset predominantly consisted of advanced stage cases as well with 79.2% of stage III and IV subjects, while the remaining datasets demonstrated a more balanced distribution across tumour stages. In terms of tumour region, the UMM3 and HN5000 datasets had a limited number of cases involving the nasopharynx, salivary glands, nasal cavity, and paranasal sinuses. In addition, the UMM1 and UMM2 datasets contained cases with tumours overlapping multiple areas. In the integrated data, the three biggest tumour site groups were oropharynx (37.1%), larynx (31.3%) and oral cavity (23.6%). Tumour stages I, II, III and IV were distributed as 24.4%, 17.9%, 13.8% and 42.0% respectively.

Survivors had an average probability of overall survival for one year after end of treatment of 0.94 [95% CI: 0.93, 0.95] and of 0.88 [0.87, 0.89] after 2 years. For BD2Decide, the 1- and 2-years overall survival were 0.95 [0.91, 0.99] and 0.86 [0.79, 0.94] respectively, for UMM1 0.88 [0.83, 0.93] and 0.76 [0.70, 0.83], and for HN5000 0.94 [0.93, 0.95] and 0.89 [0.88, 0.90] (see Figure 2).

**Figure 2.**
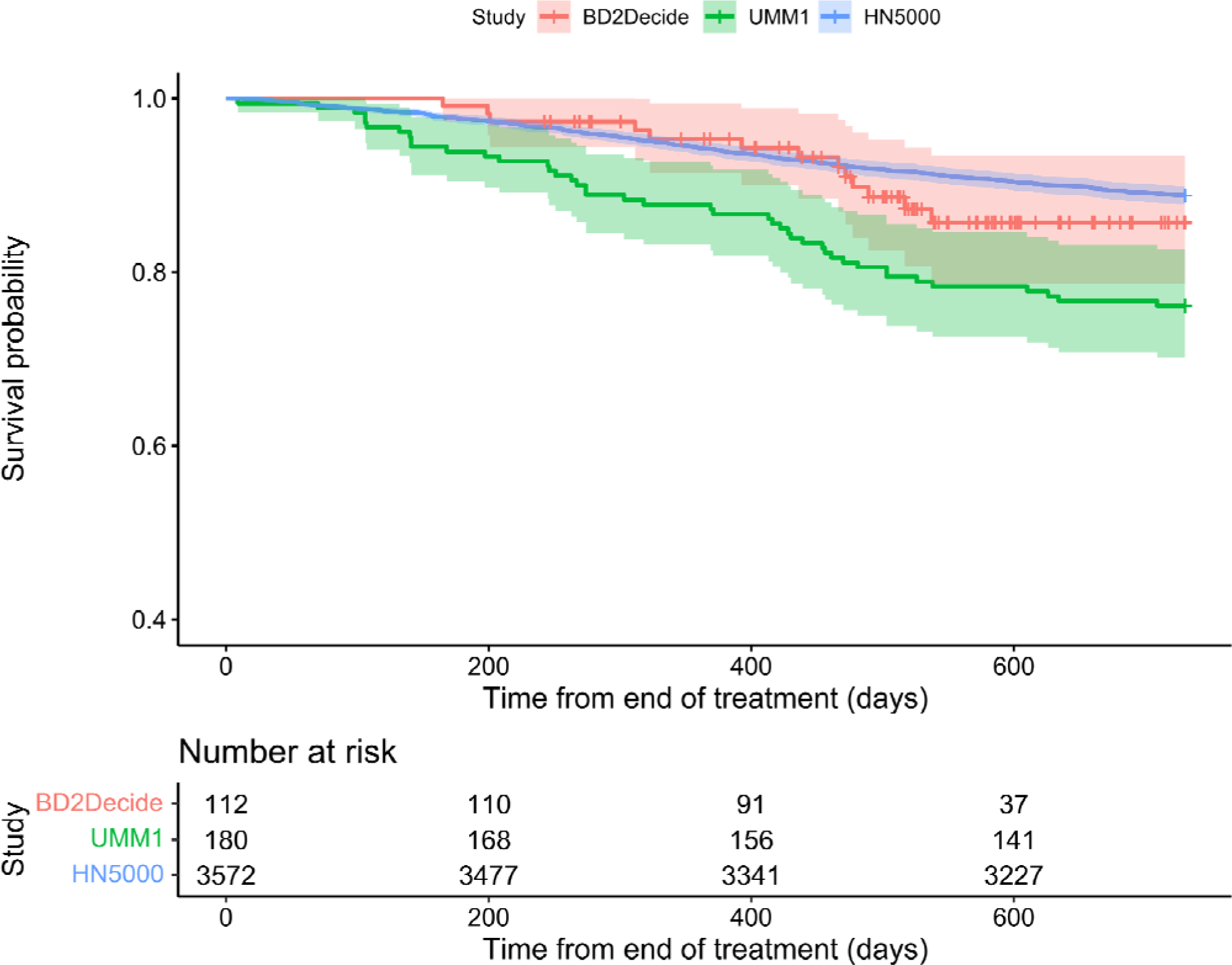
Kaplan-Meier curves with 95% confidence interval for BD2Decide, UMM1 and HN5000 studies showing 2 years overall survival after treatment. Other studies did not have the necessary time to event data for inclusion in the curve.

Concerning the GHS/QoL of survivors, we observed that survivors presented some level of QoL impairment at diagnosis and their QoL tends to decrease further during or immediately after treatment (3 and 4 months) (Figure 3). After reaching the minimum QoL, survivors recovered to a higher level (see the mean QoL at 6 and 12 months and later time points). However, according to recently established TCIs^33^ for the EORTC QLQ-C30 functioning and symptom scales, 40% and 35% of patients presented scores within ranges that indicate clinically important troubles related to physical and cognitive functioning respectively, 36 months post-diagnosis. Additionally, 30% reported fatigue and 39% indicated experiencing pain over the respective TCIs (see Table 4). Table 5 displays the data for the head and neck module, showing that the highest median scores among the EORTC QLQ-H&N35 scales were for dry mouth, coughing, sticky saliva, and reduced sexuality, with all four symptoms sharing a median value of 33.

**Figure 3.**
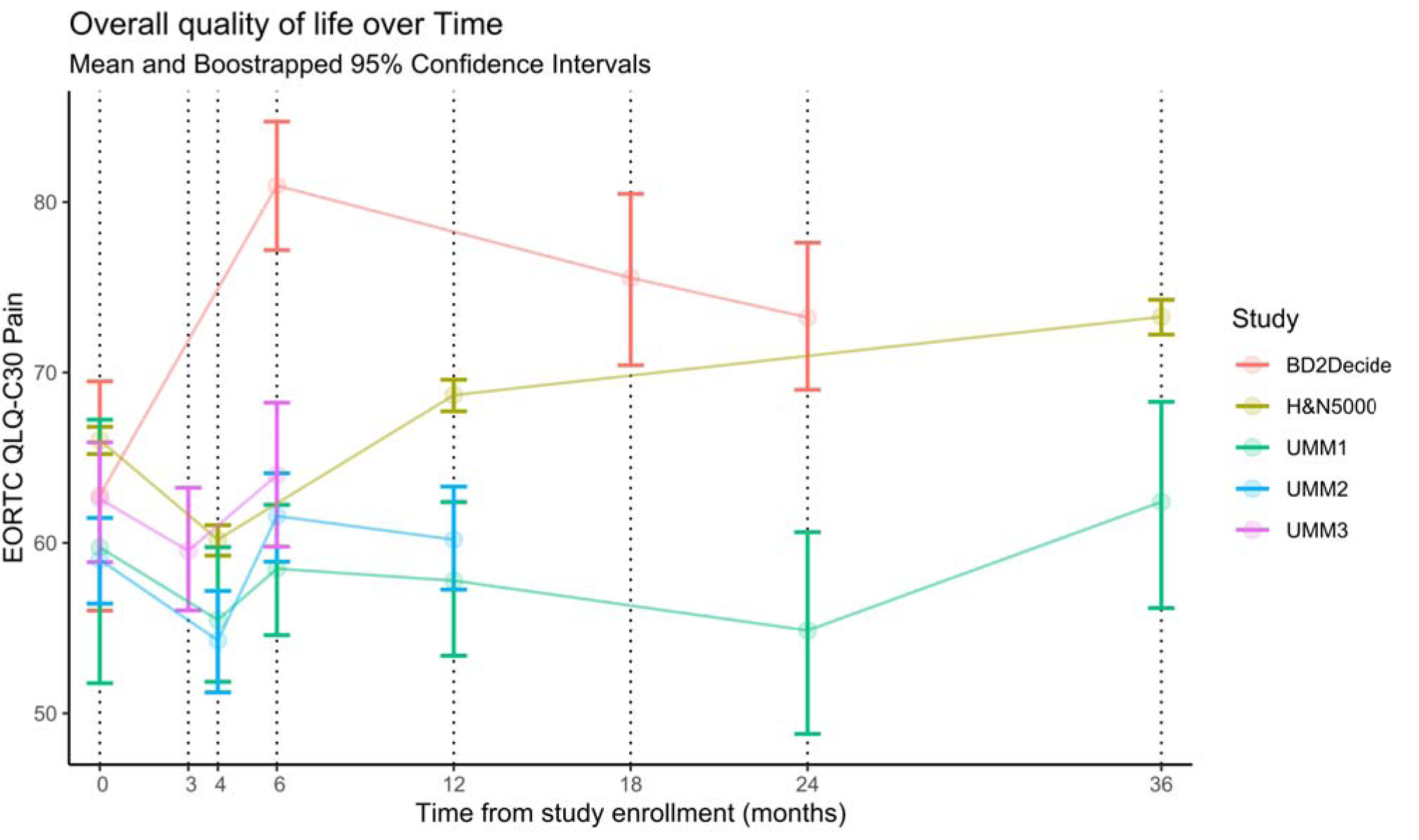
Global QoL trajectories conditional on survival and measured QoL per study. The baseline for UMM1 and UMM2 is at 4 months, UMM3 is at 3 months, BD2Decideis at 6 months, and HN5000 at 12 months. Measurements at time 0 are before treatment and all measurements before the respective baselines are for patients that survived treatment and completed the questionnaires. Patients with missing global QoL were not considered in this figure.

**Table 4.** EORTC QLQ-C30 questionnaire scales stratified by study.

| Characteristic | INT<br>N = 112<sup>1</sup> | UMM1<br>N = 180<sup>1</sup> | UMM2<br>N = 426<sup>1</sup> | UMM3<br>N = 158<sup>1</sup> | UOB<br>N = 3572<sup>1</sup> | Overall<br>N = 4448<sup>1</sup> |
| --- | --- | --- | --- | --- | --- | --- |
| Physical Functioning | 93 (86, 100) | 80 (53, 86) | 80 (66, 93) | 80 (60, 93) | 86 (66, 100) | 86 (66, 100) |
| Missing | 57 | 70 | 194 | 25 | 1,485 | 1,831 |
| Role Functioning | 100 (92, 100) | 66 (33, 100) | 66 (50, 100) | 66 (33, 100) | 83 (66, 100) | 83 (66, 100) |
| Missing | 57 | 71 | 195 | 25 | 1,471 | 1,819 |
| Cognitive Functioning | 100 (83, 100) | 100 (66, 100) | 100 (66, 100) | 100 (83, 100) | 83 (66, 100) | 83 (66, 100) |
| Missing | 57 | 70 | 194 | 29 | 1,448 | 1,798 |
| Emotional Functioning | 91 (75, 100) | 75 (50, 91) | 75 (50, 91) | 75 (58, 91) | 83 (66, 100) | 83 (66, 100) |
| Missing | 57 | 70 | 194 | 29 | 1,464 | 1,814 |
| Social Functioning | 100 (100, 100) | 66 (50, 100) | 83 (50, 100) | 83 (50, 100) | 83 (66, 100) | 83 (66, 100) |
| Missing | 57 | 70 | 194 | 28 | 1,457 | 1,806 |
| Fatigue | 11 (0, 33) | 33 (11, 55) | 33 (11, 55) | 33 (22, 55) | 33 (11, 44) | 33 (11, 44) |
| Missing | 57 | 70 | 195 | 25 | 1,454 | 1,801 |
| Pain | 0 (0, 16) | 33 (0, 50) | 16 (0, 37) | 33 (0, 50) | 16 (0, 33) | 16 (0, 33) |
| Missing | 57 | 70 | 194 | 26 | 1,461 | 1,808 |
| Nausea/Vomiting | 0 (0, 0) | 0 (0, 0) | 0 (0, 0) | 0 (0, 16) | 0 (0, 0) | 0 (0, 0) |
| Missing | 57 | 70 | 195 | 25 | 1,449 | 1,796 |
| Diarrhoea | 0 (0, 0) | 0 (0, 0) | 0 (0, 0) | 0 (0, 0) | 0 (0, 0) | 0 (0, 0) |
| Missing | 57 | 71 | 194 | 29 | 1,443 | 1,794 |
| Constipation | NA (NA, NA) | 0 (0, 0) | 0 (0, 0) | 0 (0, 33) | 0 (0, 33) | 0 (0, 33) |
| Missing | 112 | 70 | 194 | 30 | 1,442 | 1,848 |
| Appetite Loss | NA (NA, NA) | 0 (0, 33) | 0 (0, 33) | 33 (0, 66) | 0 (0, 33) | 0 (0, 33) |
| Missing | 112 | 70 | 196 | 25 | 1,456 | 1,859 |
| Insomnia | NA (NA, NA) | 0 (0, 66) | 33 (0, 66) | 33 (0, 66) | 33 (0, 33) | 33 (0, 33) |
| Missing | 112 | 70 | 195 | 26 | 1,442 | 1,845 |
| Dyspnoea | 0 (0, 33) | 33 (0, 66) | 33 (0, 33) | 0 (0, 33) | 0 (0, 33) | 0 (0, 33) |
| Missing | 57 | 70 | 195 | 25 | 1,447 | 1,794 |
| Financial Impact | 0 (0, 0) | 33 (0, 66) | 33 (0, 66) | 0 (0, 33) | 0 (0, 33) | 0 (0, 33) |
| Missing | 57 | 70 | 196 | 29 | 1,447 | 1,799 |
| GHS/QoL | 83 (75, 87) | 50 (41, 75) | 50 (33, 66) | 58 (50, 75) | 75 (58, 83) | 66 (50, 83) |
| Missing | 57 | 65 | 179 | 29 | 1,444 | 1,774 |
| GHS/QoL | 83 (75, 87) | 50 (41, 75) | 50 (33, 66) | 58 (50, 75) | 75 (58, 83) | 66 (50, 83) |
| Missing | 57 | 65 | 179 | 29 | 1,444 | 1,774 |

<sup>1</sup>Median (IQR)

**Table 5.** EORTC QLQ-H&N35 module scales stratified by study.

| **Characteristic** | **INT**<br>N = 112<sup>1</sup> | **UMM1**<br>N = 180<sup>1</sup> | **UMM2**<br>N = 426<sup>1</sup> | **UMM3**<br>N = 158<sup>1</sup> | **UOB**<br>N = 3572<sup>1</sup> | **Overall**<br>N = 4448<sup>1</sup> |
| --- | --- | --- | --- | --- | --- | --- |
| Pain | 8 (0, 25) | 16 (0, 50) | 16 (0, 41) | 16 (8, 35) | 8 (0, 25) | 16 (0, 33) |
| Missing | 60 | 61 | 181 | 26 | 1,502 | 1,830 |
| Swallowing Problems | 8 (0, 16) | 16 (0, 41) | 8 (0, 46) | 0 (0, 33) | 8 (0, 25) | 8 (0, 25) |
| Missing | 60 | 63 | 183 | 30 | 1,484 | 1,820 |
| Senses Problems | 16 (0, 33) | 66 (50, 100) | 0 (0, 16) | 33 (0, 54) | 16 (0, 50) | 16 (0, 50) |
| Missing | 60 | 61 | 178 | 26 | 1,453 | 1,778 |
| Speech Problems | 11 (0, 22) | 66 (36, 77) | 55 (33, 88) | 26 (13, 46) | 11 (0, 33) | 22 (0, 44) |
| Missing | 60 | 66 | 179 | 31 | 1,479 | 1,815 |
| Trouble with social eating | 8 (0, 16) | 25 (6, 50) | 0 (0, 25) | 25 (4, 50) | 16 (0, 41) | 16 (0, 41) |
| Missing | 60 | 64 | 183 | 31 | 1,491 | 1,829 |
| Trouble with social contact | 0 (0, 6) | 13 (0, 40) | 6 (0, 20) | 0 (0, 33) | 0 (0, 13) | 0 (0, 20) |
| Missing | 60 | 60 | 179 | 33 | 1,486 | 1,818 |
| Less Sexuality | 0 (0, 33) | 33 (0, 66) | 33 (0, 66) | 16 (0, 66) | 33 (0, 66) | 33 (0, 66) |
| Missing | 60 | 73 | 196 | 39 | 1,696 | 2,064 |
| Teeth Problems | 0 (0, 33) | 0 (0, 33) | 0 (0, 0) | 22 (0, 44) | 0 (0, 33) | 0 (0, 33) |
| Missing | 60 | 63 | 178 | 27 | 1,486 | 1,814 |
| Opening Mouth | 0 (0, 33) | 0 (0, 66) | 0 (0, 0) | 0 (0, 33) | 0 (0, 33) | 0 (0, 33) |
| Missing | 60 | 61 | 179 | 27 | 1,447 | 1,774 |
| Dry Mouth | 33 (33, 66) | 33 (0, 66) | 100 (0, 100) | 33 (16, 66) | 33 (33, 92) | 33 (33, 100) |
| Missing | 60 | 62 | 179 | 26 | 1,446 | 1,773 |
| Sticky Saliva | 33 (33, 66) | 33 (0, 66) | 100 (0, 200) | 2 (1, 3) | 33 (0, 66) | 33 (0, 66) |
| Missing | 60 | 62 | 180 | 27 | 1,458 | 1,787 |
| Coughing | 0 (0, 33) | 66 (33, 74) | 100 (100, 200) | 0 (0, 33) | 33 (0, 33) | 33 (0, 66) |
| Missing | 60 | 60 | 180 | 26 | 1,449 | 1,775 |
| Felt Ill | 0 (0, 0) | 33 (0, 66) | 100 (0, 200) | NA (NA, NA) | 0 (0, 33) | 0 (0, 33) |
| Missing | 60 | 61 | 180 | 158 | 1,453 | 1,912 |
| Pain Killers | 33 (25, 33) | 0 (0, 100) | 0 (0, 100) | NA (NA, NA) | 0 (0, 100) | 0 (0, 100) |
| Missing | 60 | 58 | 178 | 158 | 1,449 | 1,903 |
| Nutritional Supplements | 33 (33, 33) | 0 (0, 0) | 0 (0, 0) | NA (NA, NA) | 0 (0, 100) | 0 (0, 100) |
| Missing | 60 | 59 | 180 | 158 | 1,454 | 1,911 |
| Feeding Tube | 33 (33, 33) | 0 (0, 100) | 0 (0, 0) | NA (NA, NA) | 0 (0, 0) | 0 (0, 0) |
| Missing | 60 | 58 | 180 | 158 | 1,458 | 1,914 |

| Characteristic | INT<br>N = 112<sup>1</sup> | UMM1<br>N = 180<sup>1</sup> | UMM2<br>N = 426<sup>1</sup> | UMM3<br>N = 158<sup>1</sup> | UOB<br>N = 3572<sup>1</sup> | Overall<br>N = 4448<sup>1</sup> |
| --- | --- | --- | --- | --- | --- | --- |
| Weight Loss | 33 (33, 33) | 0 (0, 100) | 0 (0, 100) | 0 (0, 33) | 0 (0, 0) | 0 (0, 0) |
| Missing | 60 | 60 | 180 | 29 | 1,473 | 1,802 |
| Weight Gain | 33 (33, 33) | 0 (0, 100) | 0 (0, 0) | NA (NA, NA) | 0 (0, 100) | 0 (0, 100) |
| Missing | 60 | 59 | 181 | 158 | 1,508 | 1,966 |

<sup>1</sup>Median (IQR)

### 3.2 Missing data

The proportion of missing values depended on the type of variable. Most clinical variables (e.g., sex, age, tumour stage and location and treatment) presented low missingness of 0-6.4%. Variables collected through interviews (e.g., marital status, education level and tobacco usage) had moderate missingness of 25.9-39.3%, while QoL variables had higher missingness, 39.9-41.8% at baseline. The majority of missing data in QoL variables was related with missing of the whole questionnaire (N=1692), usually because patients did not send them back, whereas 8.8% of subjects (N=393) presented missing values only in specific QoL subscales while others were measured (See Table 6 and figure S1 in the Supplementary Information). It is important to notice that the high proportion of unfilled questionnaires is driven by patients receiving the questionnaires by post and never returning them, while most patients that returned the questionnaire answered all questions. Finally, some data was missing in blocks in the dataset because studies did not collect the exact same variables. Body mass index (BMI) information was solely available for HN5000, and BD2Decide lacked data on education level and income. Furthermore, the UMM3 dataset did not include information on alcohol consumption and smoking status.

**Table 6.** Odds ratios (OR) and p-values from univariate logistic regression models with response *“Missing GHS/QoL”* given the respective covariate in the table.

|  | OR | p-value |
| --- | --- | --- |
| **Sex** |  |  |
| Male | — | — |
| Female | 0.88 | <.001 |
| **Age** | 0.99 | <.001 |
| **Education level** |  |  |
| Low | — | — |
| Medium | 0.81 | 0.02 |
| High | 0.51 | <.001 |
| **Income** |  |  |
| Less than 500 | — | — |
| 500 - 1500 | 0.74 | 0.02 |
| 1501 - 2500 | 0.64 | 0.001 |
| 2501 - 3500 | 0.65 | 0.02 |
| More than 3500 | 0.50 | <.001 |
| **Body mass index** | 0.99 | 0.05 |
| **Marital status** |  |  |
| Single | — | — |
| Married/Living with a partner | 0.74 | 0.01 |
| Divorced/Separated | 1.07 | 0.62 |
| Widowed | 0.82 | 0.28 |
| **Smoker status at the baseline** |  |  |
| Current | — | — |
| Former | 0.55 | <.001 |
| Never | 0.48 | <.001 |
| **Units of alcohol per month** | 1.00 | 0.002 |
| **Tumour stage (TNM 7<sup>th</sup> Ed)** |  |  |
| I | — | — |
| II | 1.20 | 0.04 |
| III | 1.00 | 0.89 |
| IV | 1.11 | 0.15 |
| **Tumour region** |  |  |
| Oral cavity | — | — |
| Oropharynx | 0.94 | 0.47 |
| **Nasopharynx** | 0.87 | 0.58 |
| **Hypopharynx** | 1.10 | 0.28 |
| **Larynx** | 1.09 | 0.28 |
| **Overlapping several areas** | 1.27 | 0.50 |
| **Nasal cavity and paranasal sinuses** | 1.05 | 0.86 |
| **Radiotherapy** |  |  |
| **Yes** | — |  |
| **No** | 0.80 | 0.002 |
| **Chemotherapy** |  |  |
| **Yes** | — | — |
| **No** | 0.94 | 0.31 |
| **Surgery** |  |  |
| **Yes** | — | — |
| **No** | 0.99 | 0.83 |

There were several associations between the frequency of missing QoL measurements and certain patient features (p-value < .05, Table 6). Specifically, being a female (OR: 0.88), older age (OR: 0.99), possessing a high level of education (OR: 0.48), having a high income (OR: 0.51), never being a smoker (OR: 0.48), being married or living with a partner (OR: 0.74), and treatment without radiotherapy (OR: 0.80) were found to be associated with a lower frequency of missing QoL data (see Table 6 for the full list of odds ratios).

## 4 Discussion

In this study, we defined and built a cohort of HNC survivors from historical data contributed from studies conducted in Germany, Italy, and the UK, with clear eligibility criteria and survivorship definition. To the best of our knowledge this is the largest cohort of HNC survivors in the world.

We observed large interstudy heterogeneity in global health status GHS/QoL at the survivorship baseline, i.e., at the first QoL measurement after end of treatment. Two important factors that can impact GHS/QoL should be considered: the time at which the baseline GHS/QoL was measured and the amount of missingness. Usually, QoL immediately after treatment is lower than at later timepoints^36^, which can explain the lower GHS/QoL in some contributing studies compared to others. For example, the UMM studies measured QoL just after treatment completion (3-4 months after diagnosis), while the HN5000 measured QoL 12 months after diagnosis, which may be several months after actual end of treatment for some patients. BD2Decide measured QoL 6 months after diagnosis, but the proportion of missing values is 50.9%. Since the missingness is related to the measurement, e.g., patients with the lowest GHS/QoL might not answer the questionnaire, the bias introduced can lead to a higher QoL on average than expected.

Overall survival 1-2 years after treatment remained high on average, but there were differences between the studies with UMM1 having shorter survival than BD2Decide and HN5000. This may be related to the patient characteristics in the studies but may also be partly explained by how end-of-treatment (baseline) was defined in the studies. In UMM1 the baseline was at 4 months after diagnosis, while the baseline was later in BD2Decide (at 6 months after diagnosis) and in HN5000 (12 months after diagnosis).

Regarding the QoL trajectory over time, we observe a dip in GHS/QoL around treatment, followed by an increase after treatment and a flattening out over time. This pattern has also been observed in previous studies^37–39^. Based on the EORTC QLQ-H&N35 scales, the most prominent concerns for this cohort are related to physical symptoms such as dry mouth, sticky saliva, and coughing.

We identified being female, older age, high education level, high income, and not smoking as factors associated with lower probability of not completing the GHS/QoL scale. This highlights the importance of proper missing data handling for improving the interpretability and analysis of QoL data in this population, as complete case approaches can introduce selection bias and impact the validity of the results. Furthermore, it is important to acknowledge the presence of data missing in blocks in the dataset, defined as structured missingness (SM)^40^. SM can be caused by several reasons, but it arises naturally when integrating multisource data due to studies collecting different sets of variables to address different research questions. This is a missing pattern that is becoming more common due to the increase in use of multisource data integration for machine learning models development. In the BD4QoL data SM is also present due to patients who never received the questionnaires or never returned them by post.

The BD4QoL historical cohort has several important strengths. It is to date the largest dataset on HNC survivors with 4448 subjects. The BD4QoL study is composed mainly by HNC survivors from 3 countries in Europe (Italy, Germany, and UK), in addition to smaller groups from many countries in different continents (Americas, Asia and Europe) included in the UMM3 study. This provides a dataset with rich demographics and regional variations. The data contains repeated QoL measurements acquired at various timepoints giving opportunity to unveil the longitudinal course of QoL in HNC survivors. The standardized and validated EORTC QLQ-C30 and the head and neck modules H&N35/43 were applied in all studies integrated in the BD4QoL cohort, which provides rich information about multidomain QoL. This can be used to describe HNC survivors, identify their needs to improve patient care and long-term QoL. The data providers maintained high quality data standards in the original studies, which consequently make the integrated BD4QoL data of excellent quality as well.

Nonetheless, it is important to state weaknesses in this study that can limit the scope of potential future analyses. Most of the survivors in the BD4QoL historical cohort are white Europeans, with the Global South and other ethnicities being underrepresented – though this represents the population where the studies were conducted. The number of questionnaires that were not returned by the patients and the patterns of missing data identified have the potential to introduce biases in future analyses, so missing data handling strategies should be carefully considered. In addition, structured missingness is present which requires specialized methods if the affected variables are kept in the analyses^41,42^. The heterogeneity in the time of which patients are considered survivors – whether after 3, 4, 6, or 12 months – is also a challenging aspect in this data. All patients are survivors, but those who have survived for longer, like 12 months, may not only have different QoL but also a greater likelihood of surviving even longer.

## 5 Findings to date

Time is often a neglected variable in clinical studies, but it has a clear impact in PROs^43^. Instruments measuring PROs are administered at various timepoints across studies due to the different research questions and assumed intervention effects. Quality of life measurements are sensitive to time of collection and impose a challenge for interoperability when different datasets are to be integrated.

However, measuring QoL “as often as possible”, which is sometimes done, is also not a solution for this problem because study participants then lose the motivation to complete the forms properly and completely. As a rule of thumb, about 4 times a year in the first year after diagnosis and once a year after that are usually acceptable by patients.

## 6 Ethics statement

Our study was conducted in full accordance with ethical principles, including the World Medical Association’s Declaration of Helsinki (2002 version). The study protocol received ethical approval by the Norwegian Regional Committee for Medical and Health Research Ethics (REK) South-East D under application number 154191. The data is stored in compliance with GDPR legislation in the secure server for sensitive data at the University of Oslo (TSD/USIT) and access is granted to authorised collaborators included in the ethical approval. Each individual study that provided data received ethical approval from local authorities in Italy, Germany, the UK, and all further countries involved. The data providers submitted copies of these ethical approvals to the principal investigator.

BD2Decide study was approved by institutional research ethics board with identifier N. INT 65/16.

The HN5000 study was approved by the National Research Ethics Committee (South West Frenchay Ethics Committee, reference number 10/H0107/57, 5 November 2010) and the Research and Development departments of participating NHS Trusts. Informed consent was obtained from all patients recruited to HN5000.

UMM1 and UMM2 received ethical approval from the Leipzig University Ethics Committee. UMM3 was approved by Landesa rztekammer Rheinland-Pfalz ethics committee under approval number 837.281.14 (9520).

## 8 Author Contributions

MMS, KT, SS, KH, ST, AN, CV, LL-P, MFC-U, GF, LL, ML contributed to the conceptualization of the study. SS, KH, ST, AN, MFC-U, AF, LL, ML were responsible for funding acquisition. KT, SS, KH, ST, MP, AN, SC, LL conducted data sharing. MMS, EIFF, AF, ML participated in the investigation process. MMS, EIFF developed and/or worked with the necessary software. MMS, EIFF, KT, CV, ML crafted the methodology framework of the study. LL-P, AF, LL provided the resources needed for the research. All authors contributed to writing the manuscript.

## 10 Acknowledgments

This work was performed on the TSD (Tjeneste for Sensitive Data) facilities, owned by the University of Oslo, operated and developed by the TSD service group at the University of Oslo, IT-Department (USIT). The statistical analysis and data harmonisation were performed on resources provided by Sigma2 - the National Infrastructure for High Performance Computing and Data Storage in Norway. The Authors thank the researchers and clinicians who designed the BD2Decide, HN5000, and Mainz studies, the research, laboratory and clinical staff who supported the conduct of the studies; and the people with head and neck cancer who took part.

## 11 Data Availability Statement

The BD4QoL historical cohort dataset is hosted by the Services for Sensitive Data (TSD) at the University of Oslo. Access to the data may be granted by the data owners upon application.

The data that support the findings of this study are available from head and neck 5000. Further information may be found on the Head and Neck 5000 website: https://www.headandneck5000.org.uk/information-for-researchers.

## Footnotes

1 Alcohol unit is a dimensionless measurement unit defined as 10ml or 8g of pure alcohol.

## References

1. Mody MD, Rocco JW, Yom SS, Haddad RI, Saba NF. Head and neck cancer. The Lancet. 2021;398(10318):2289–2299. doi:10.1016/S0140-6736(21)01550-6

2. Vos T, Abajobir AA, Abate KH, et al. Global, regional, and national incidence, prevalence, and years lived with disability for 328 diseases and injuries for 195 countries, 1990–2016: a systematic analysis for the Global Burden of Disease Study 2016. The Lancet. 2017;390(10100):1211–1259. doi:10.1016/S0140-6736(17)32154-2

3. Mehanna H, Paleri V, West CML, Nutting C. Head and neck cancer--Part 1: Epidemiology, presentation, and prevention. BMJ. 2010;341(sep20 1):c4684–c4684. doi:10.1136/bmj.c4684

4. Ferlay J, Colombet M, Soerjomataram I, et al. Cancer statistics for the year 2020: An overview. Int J Cancer. 2021;149(4):778–789. doi:10.1002/ijc.33588

5. Pulte D, Brenner H. Changes in Survival in Head and Neck Cancers in the Late 20th and Early 21st Century: A Period Analysis. The Oncologist. 2010;15(9):994–1001. doi:10.1634/theoncologist.2009-0289

6. Høxbroe Michaelsen S, Grønhøj C, Høxbroe Michaelsen J, Friborg J, von Buchwald C. Quality of life in survivors of oropharyngeal cancer: A systematic review and meta-analysis of 1366 patients. European Journal of Cancer. 2017;78:91–102. doi:10.1016/j.ejca.2017.03.006

7. Ramqvist T, Dalianis T. Oropharyngeal Cancer Epidemic and Human Papillomavirus. Emerg Infect Dis. 2010;16(11):1671–1677. doi:10.3201/eid1611.100452

8. Carlander ALF, Grønhøj Larsen C, Jensen DH, et al. Continuing rise in oropharyngeal cancer in a high HPV prevalence area: A Danish population-based study from 2011 to 2014. European Journal of Cancer. 2017;70:75–82. doi:10.1016/j.ejca.2016.10.015

9. Ang KK, Harris J, Wheeler R, et al. Human Papillomavirus and Survival of Patients with Oropharyngeal Cancer. N Engl J Med. 2010;363(1):24–35. doi:10.1056/NEJMoa0912217

10. Patel MA, Blackford AL, Rettig EM, Richmon JD, Eisele DW, Fakhry C. Rising population of survivors of oral squamous cell cancer in the United States: Rising Oral Cancer Survivors in the US. Cancer. 2016;122(9):1380–1387. doi:10.1002/cncr.29921

11. Goyal N, Day A, Epstein J, et al. Head and neck cancer survivorship consensus statement from the American Head and Neck Society. Laryngoscope Investig Oto. 2022;7(1):70–92. doi:10.1002/lio2.702

12. Forastiere A, Koch W, Trotti A, Sidransky D. Head and Neck Cancer. N Engl J Med. 2001;345(26):1890–1900. doi:10.1056/NEJMra001375

13. Vermorken JB, Specenier P. Optimal treatment for recurrent/metastatic head and neck cancer. Annals of Oncology. 2010;21:vii252–vii261. doi:10.1093/annonc/mdq453

14. Strojan P, Hutcheson KA, Eisbruch A, et al. Treatment of late sequelae after radiotherapy for head and neck cancer. Cancer Treatment Reviews. 2017;59:79–92. doi:10.1016/j.ctrv.2017.07.003

15. Alterio D, Jereczek-Fossa BA, Franchi B, et al. Thyroid disorders in patients treated with radiotherapy for head-and-neck cancer: A retrospective analysis of seventy-three patients. International Journal of Radiation Oncology*Biology*Physics. 2007;67(1):144–150. doi:10.1016/j.ijrobp.2006.08.051

16. Taylor K, Krüger M, Singer S. Long-term toxicity among head and neck cancer patients—A systematic review. Onkologe. 2021;27(S2):145–149. doi:10.1007/s00761-021-00914-x

17. Quinten C, Martinelli F, Coens C, et al. A global analysis of multitrial data investigating quality of life and symptoms as prognostic factors for survival in different tumor sites. Cancer. 2014;120(2):302–311. doi:10.1002/cncr.28382

18. van Nieuwenhuizen AJ, Buffart LM, Brug J, René Leemans C, Verdonck-de Leeuw IM. The association between health related quality of life and survival in patients with head and neck cancer: A systematic review. Oral Oncology. 2015;51(1):1–11. doi:10.1016/j.oraloncology.2014.09.002

19. So W, Chan R, Chan D, et al. Quality-of-life among head and neck cancer survivors at one year after treatment–a systematic review. European journal of cancer. 2012;48(15):2391–2408.

20. Taylor KJ, Amdal CD, Bjordal K, et al. Serious Long-Term Effects of Head and Neck Cancer from the Survivors’ Point of View. Healthcare. 2023;11(6):906. doi:10.3390/healthcare11060906

21. Alonso I, Lopez-Perez L, Guirado JCM, Cabrera-Umpierrez MF, Arredondo MT, Fico G. Data analytics for predicting quality of life changes in head and neck cancer survivors: a scoping review. In: 2021 43rd Annual International Conference of the IEEE Engineering in Medicine & Biology Society (EMBC). IEEE; 2021:2262–2265.

22. Taylor K, Singer S. Long-term quality of life in head and neck cancer patients: A systematic review. Onkologe. 2019;25(S2):125–131. doi:10.1007/s00761-019-0527-z

23. Aaronson NK, Ahmedzai S, Bergman B, et al. The European Organization for Research and Treatment of Cancer QLQ-C30: A Quality-of-Life Instrument for Use in International Clinical Trials in Oncology. JNCI Journal of the National Cancer Institute. 1993;85(5):365–376. doi:10.1093/jnci/85.5.365

24. Fayers P, Aaronson NK, Bjordal K, Sullivan M. EORTC QLQ–C30 Scoring Manual. European Organisation for Research and Treatment of Cancer; 1995.

25. The Head and Neck 5000 Study Team, Ness AR, Waylen A, et al. Establishing a large prospective clinical cohort in people with head and neck cancer as a biomedical resource: head and neck 5000. BMC Cancer. 2014;14(1):973. doi:10.1186/1471-2407-14-973

26. Ness AR, Waylen A, Hurley K, et al. Recruitment, response rates and characteristics of 5511 people enrolled in a prospective clinical cohort study: head and neck 5000. Clinical Otolaryngology. 2016;41(6):804–809. 10.1111/coa.12548

27. Singer S, Danker H, Guntinas–Lichius O, et al. Quality of life before and after total laryngectomy: Results of a multicenter prospective cohort study. Head & Neck. 2014;36(3):359–368. doi:10.1002/hed.23305

28. Clasen D, Keszte J, Dietz A, et al. Quality of life during the first year after partial laryngectomy: Longitudinal study. Head & Neck. 2018;40(6):1185–1195. doi:10.1002/hed.25095

29. Singer S, Amdal CD, Hammerlid E, et al. International validation of the revised European Organisation for Research and Treatment of Cancer Head and Neck Cancer Module, the EORTC QLQ_JHN43: Phase IV. Head & Neck. 2019;41(6):1725–1737. doi:10.1002/hed.25609

30. Cavalieri S, De Cecco L, Brakenhoff RH, et al. Development of a multiomics database for personalized prognostic forecasting in head and neck cancer: The Big Data to Decide EU Project. Head & neck. 2021;43(2):601–612.

31. Compton SEC, Cancer AJC on, others. the 7th edition of the AJCC cancer staging manual and the future of TNM Ann. Surg Oncol. 17:1471–1474.

32. Almeida A, Bilbao-Jayo A, Hernandez L, et al. An Ontology for Quality of Life Modeling in Head and Neck Cancer. In: 2022 7th International Conference on Smart and Sustainable Technologies (SpliTech). IEEE; 2022:1–5. doi:10.23919/SpliTech55088.2022.9854379

33. Giesinger JM, Loth FLC, Aaronson NK, et al. Thresholds for clinical importance were established to improve interpretation of the EORTC QLQ-C30 in clinical practice and research. Journal of Clinical Epidemiology. 2020;118:1–8. doi:10.1016/j.jclinepi.2019.10.003

34. Lopez-Perez L, Hernández L, Ottaviano M, et al. BD2Decide: Big Data and Models for Personalized Head and Neck Cancer Decision Support. In: 2019 IEEE 32nd International Symposium on Computer-Based Medical Systems (CBMS). ; 2019:67–68. doi:10.1109/CBMS.2019.00024

35. Hernández L, Estévez-Priego E, López-Pérez L, et al. HeNeCOn: An ontology for integrative research in Head and Neck cancer. International Journal of Medical Informatics. 2024;181:105284. doi:10.1016/j.ijmedinf.2023.105284

36. Sandstrom, RN, MSN, APRN-BC, AOCN SK, R. Mazanec, PhD, RN, AOCN S, Gittleman, MS H, S. Barnholtz-Sloan, PhD J, Tamburro, LISW-S N, J. Daly, PhD, RN, FAAN B. A Descriptive, Longitudinal Study of Quality of Life and Perceived Health Needs in Patients With Head and Neck Cancer. JADPRO. 2016;7(6). doi:10.6004/jadpro.2016.7.6.6

37. de Vries J, Bras L, Sidorenkov G, et al. Frailty is associated with decline in health-related quality of life of patients treated for head and neck cancer. Oral Oncology. 2020;111:105020. doi:10.1016/j.oraloncology.2020.105020

38. Abel E, Silander E, Nordström F, et al. Fatigue in Patients With Head and Neck Cancer Treated With Radiation Therapy: A Prospective Study of Patient-Reported Outcomes and Their Association With Radiation Dose to the Cerebellum. Advances in Radiation Oncology. 2022;7(5):100960. doi:10.1016/j.adro.2022.100960

39. Braam PM, Roesink JM, Raaijmakers CP, Busschers WB, Terhaard CH. Quality of life and salivary output in patients with head-and-neck cancer five years after radiotherapy. Radiat Oncol. 2007;2(1):3. doi:10.1186/1748-717X-2-3

40. Mitra R, McGough SF, Chakraborti T, et al. Learning from data with structured missingness. Nat Mach Intell. 2023;5(1):13–23. doi:10.1038/s42256-022-00596-z

41. Kamphuis R, Jolani S, Lugtig P. The Blocked Imputation Approach for Missing Data. Published online 2018. doi:10.13140/RG.2.2.12467.32803

42. Tierney NJ, Harden FA, Harden MJ, Mengersen KL. Using decision trees to understand structure in missing data. BMJ Open. 2015;5(6):e007450. doi:10.1136/bmjopen-2014-007450

43. J. Pater, D. Osoba, B. Zee, et al. Effects of Altering the Time of Administration and the Time Frame of Quality of life Assessments in Clinical Trials: An Example Using the EORTC QLQ-C30 in a Large Anti-Emetic Trial. Quality of Life Research. 1998;(7):273–278. doi:10.1023/A:1024954518241

