## Supplementary Material for "Quality of life in head and neck cancer survivors: the Big Data for Quality of Life study"

### 1 Supplementary Material

#### 1.1 Data harmonization

##### Differences and commonalities in the studies variables

We observed high heterogeneity with respect to variables collected across studies. For instance, the UMM data contain a broad range of variables with detailed information about treatment tolerance, clinical devices used, welfare status such as sick leave and EU pension applications, among many other demographic and socioeconomic indicators. On the other hand, the INT data focus mainly on clinical information. The UoB data present a mix of clinical information and QoL data with several different questionnaires. We integrated basic clinical characteristics, demographics, and two quality-of-life questionnaires that were present in all datasets, the EORTC QLQ-C30 and the head and neck module EORTC QLQ-H&N35. For variables that were not completely equivalent and presented different levels of details, e.g., education level, household income, and marital status, we reduced their granularity to allow comparable measurements.

##### Time-to-event data

The UMM2 and UMM3 studies were not intended to model survival, so only interval-censored data with right censoring are available. Patients' statuses were checked at scheduled timepoints, but times of death were not recorded. All patient statuses were measured at baseline, so left censoring is not present in the data. UMM1 provided vital information for 83% of the subjects (N=181) from study enrollment (before treatment) up to 12 years. We found no significant differences between the groups with measured and missing vital information age, sex, tumor location, tumor stage, income, and education level, using a univariate logistic regression model approach where the response variable is the binary indicator *"missing vital information"* (yes/no). Therefore, we assume that the vital information is missing completely at random (MCAR) due to the data collection procedure, which involved retrieving vital information for each patient directly from clinicians many years after the study's conclusion. INT provided the time of death/censoring from study enrollment (before treatment) up to two years. UoB provided survival data with time of death/censoring from the time of study enrollment (before treatment). Participants' statuses were checked every 6 months from UK registry data up to 6 years after study enrollment. The survival data for INT, UMM1, and UoB were aligned and shifted to the BD4QoL baseline (after treatment), which consisted of a 6-month shift for INT, 4 months for UMM1, and 12 months for UoB.

### 2 Supplementary Figures and Tables

#### 2.1 Supplementary Figures

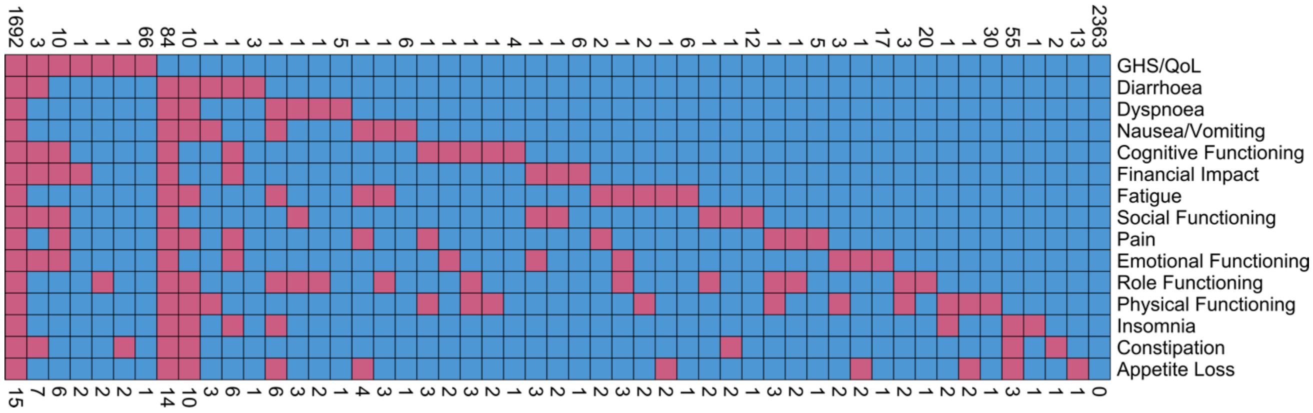


**Figure S1 – Missing patterns in the EORTC QLQ-C30 subscales at baseline. Each row identifies a variable and each column a missing pattern. The figure displays the number of cases with a given pattern in the top of each column and the respective number of missing variables in the bottom. The two biggest groups are the ones completely missing the questionnaire (N=1692) and the ones with complete questionnaire (N=2363), followed by survivors with available GHS/QoL and any other missing subscale (N=312) and survivors with missing GHS/QoL and any other subscale available (N=82).**

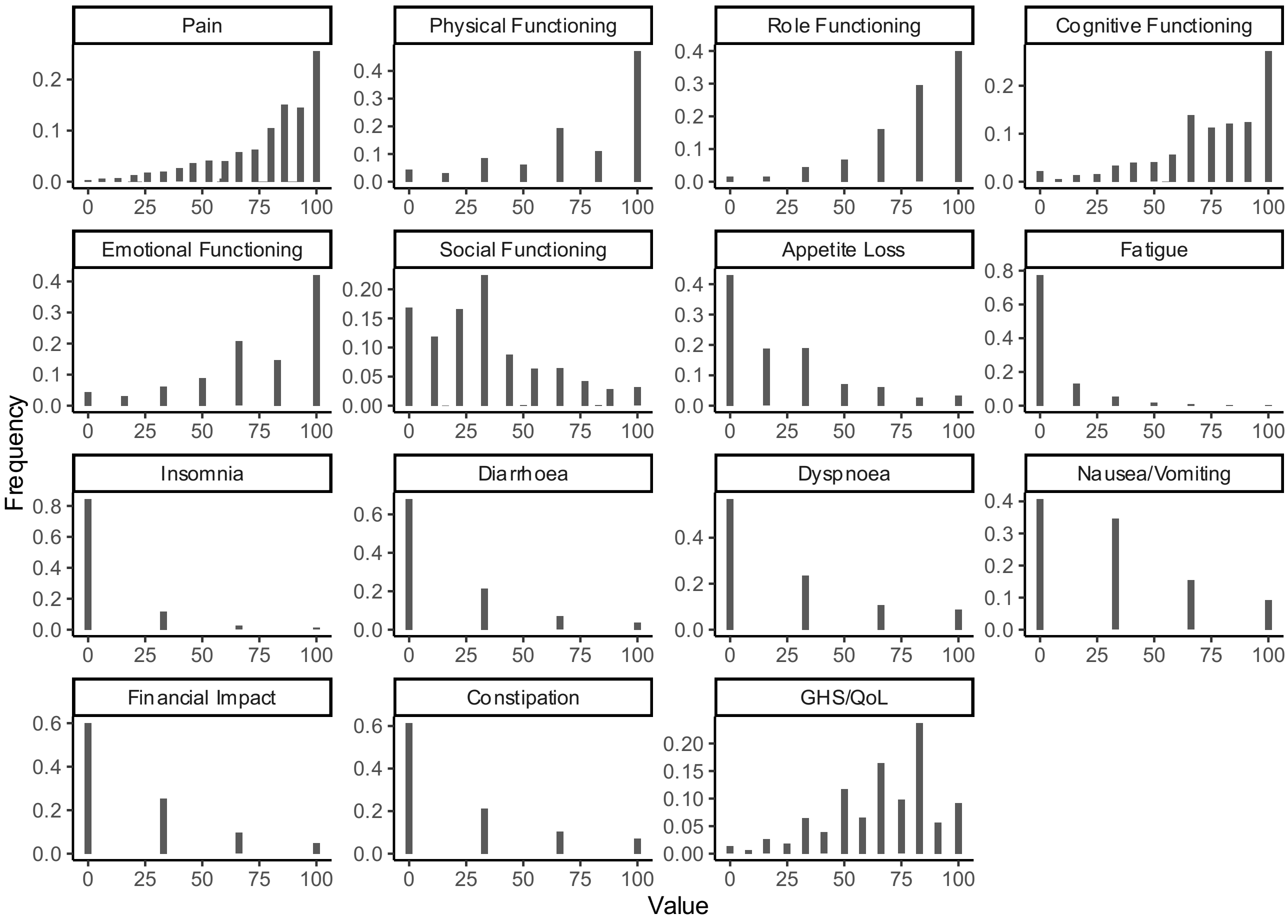


**Figure S2 EORTC QLQ-C30 functional and symptoms scales histograms.**

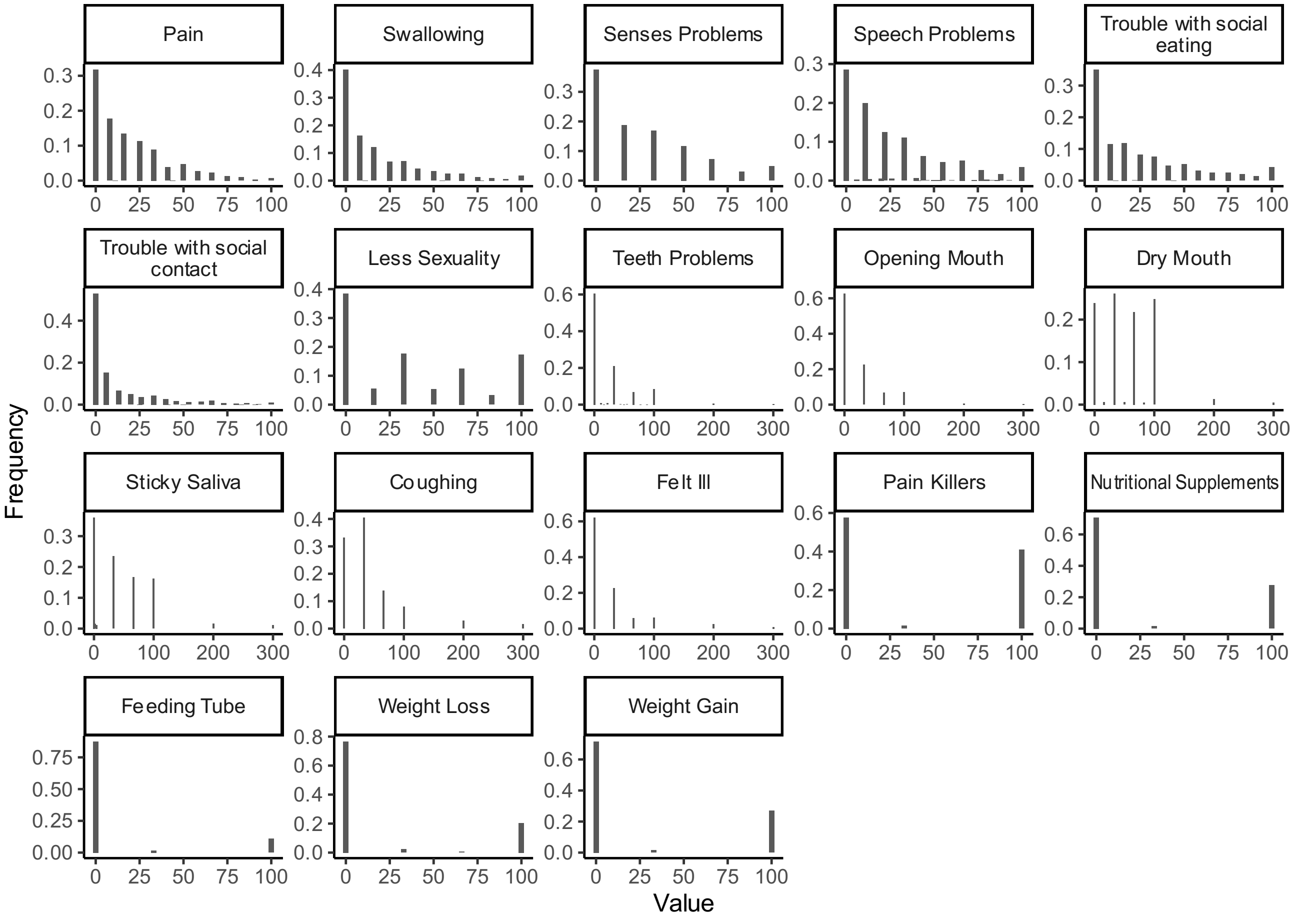


**Figure 3 - EORTC QLQ-H&N35 scales histograms.**

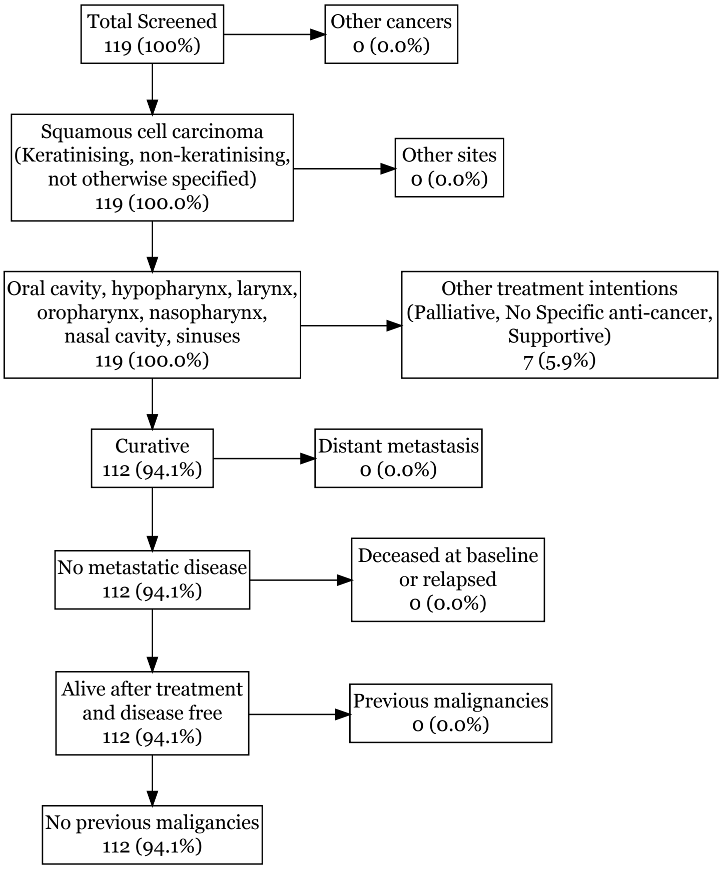


**Figure S4 – INT cohort inclusion flowchart.**

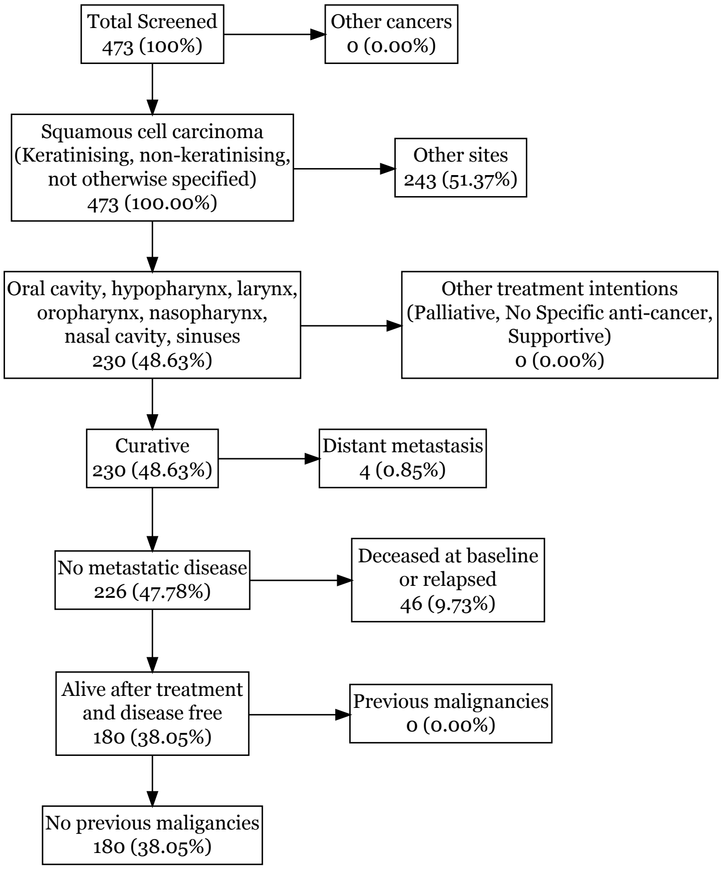


**Figure S5 – UMM1 cohort flowchart.**

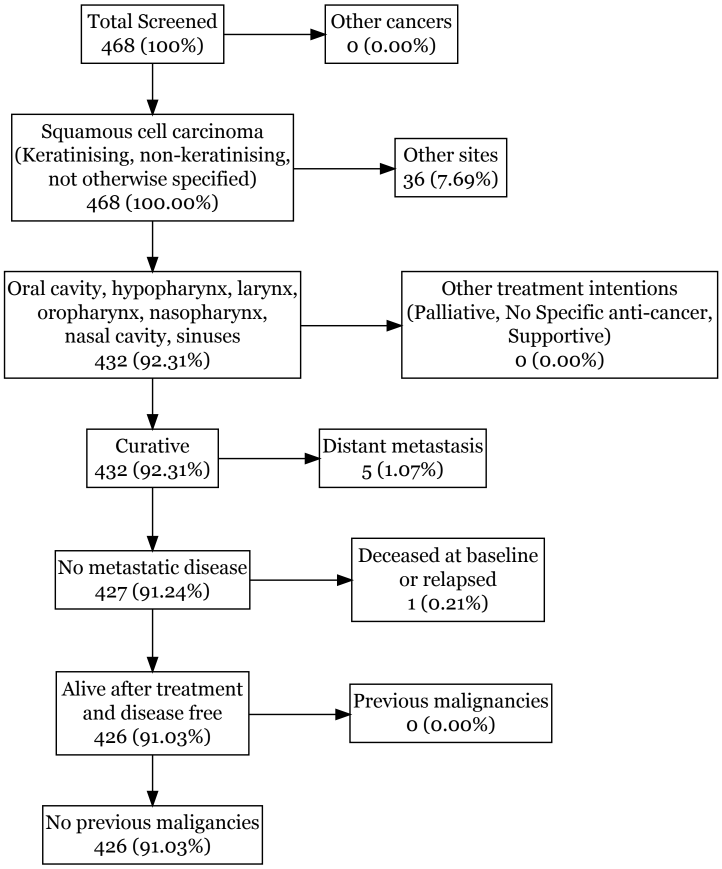


**Figure S6 - UMM2 cohort flowchart.**

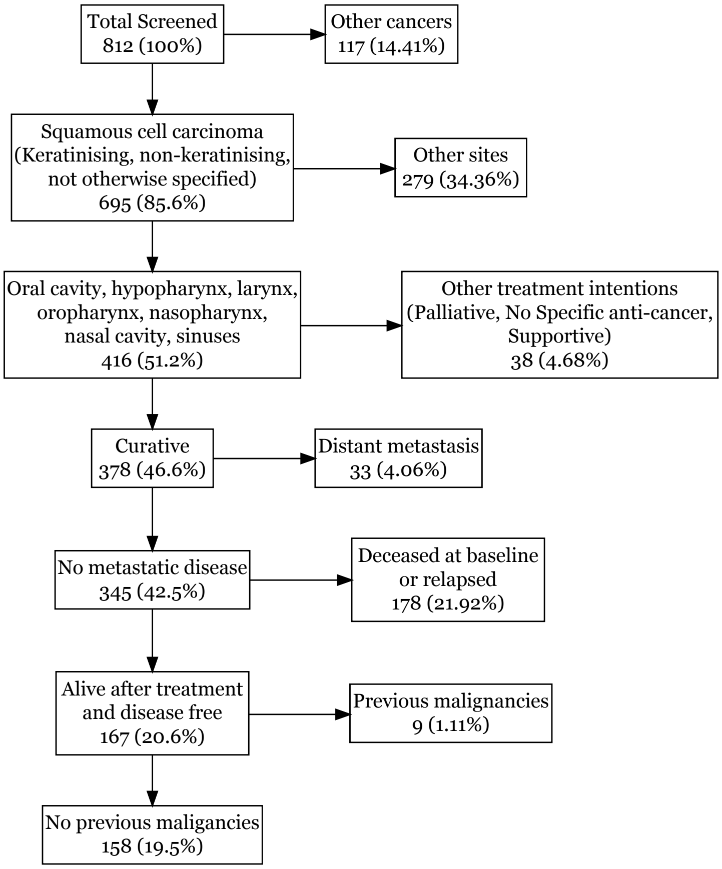


**Figure S7 - UMM3 cohort flowchart.**

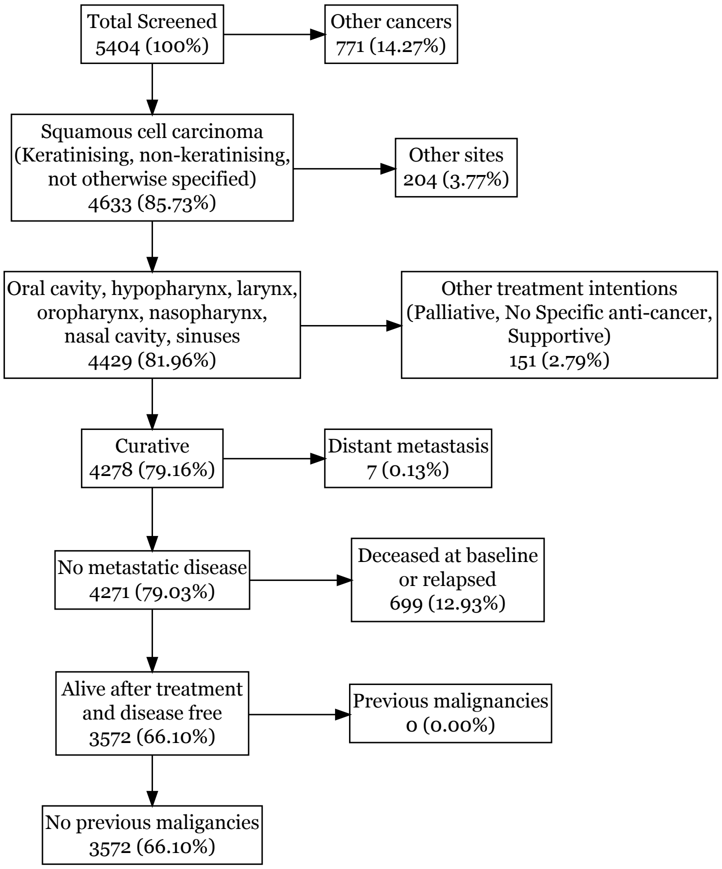


**Figure 8 - HN5000 cohort flowchart.**
